# Proton pump inhibitors and upper gastrointestinal cancer: a matched case-control study addressing confounding by indication

**DOI:** 10.1101/2025.06.13.25329558

**Authors:** Ibrahim O. Sawaid, Zohar Din, Efrat Golan, Eytan Ruppin, Avivit Golan-Cohen, Ilan Green, Eugene Merzon, Shlomo Vinker, Abraham O. Samson, Ariel Israel

**Author notes:** **Corresponding Author:** Ariel Israel, MD, PhD, Leumit Research Institute, Leumit Health Services.

## Abstract

**Objectives:** To assess the association between proton pump inhibitor (PPI) use and upper gastrointestinal (GI) cancer while addressing potential confounding by indication.

**Design:** Matched case-control study using multivariable conditional logistic regression.

**Setting:** Electronic health records from a national health provider.

**Participants:** Patients diagnosed with upper GI cancer (n=875), each matched with 10 cancer-free controls (n=8750) by age, sex, and ethnic group.

**Main outcome measures:** Adjusted odds ratios (aORs) for upper GI cancer associated with prior exposure to PPIs, H2 receptor antagonists, or antacids, with medication exposure modelled as multiple binary variables corresponding to distinct time windows before the index date. Additional models adjusted for GI-related diagnoses recorded prior to the index date (e.g., gastritis, gastroesophageal reflux disease, peptic ulcer disease).

**Results:** PPI use in the five years before the index date was initially associated with increased odds of upper GI malignancy (e.g., esomeprazole aOR 3.90 [95% CI 3.14 to 4.84]; omeprazole aOR 2.60 [2.22 to 3.06]). However, when exposure was modelled as separate binary variables for each time window, the association was strongest for use within six months of diagnosis and was not observed—or reversed—for more remote exposures. After excluding the final year before diagnosis and adjusting for symptom-related diagnoses, no positive association remained. Remote PPI use was associated with reduced risk (e.g., omeprazole more than 3 years before the index date: aOR 0.62 [0.51 to 0.75]).

**Conclusions and Relevance:** The association between PPI use and upper GI malignancy appears to reflect confounding by indication, with PPIs prescribed in response to early symptoms associated with increased cancer risk. After accounting for timing of use and underlying GI diagnoses, no harmful association remained. These findings suggest that new-onset upper GI symptoms warrant investigation for malignancy, rather than attribution of risk to acid-suppressive therapy.

## Introduction

Proton pump inhibitors (PPIs) are widely used in the treatment of acid-related gastrointestinal disorders[1]. Since the introduction of omeprazole in 1988, several PPIs— such as lansoprazole, pantoprazole, rabeprazole, esomeprazole, and dexlansoprazole[2]— have become mainstays in the management of gastroesophageal reflux disease (GERD), peptic ulcer disease, Zollinger–Ellison syndrome, and *Helicobacter pylori* infections[3]. PPIs irreversibly inhibit the gastric H^+^ /K^+^-ATPase, leading to prolonged suppression of gastric acid secretion[4].

Before the advent of PPIs, histamine-2 receptor blockers (H2RBs), such as famotidine and ranitidine, were the primary agents for acid suppression[5]. However, the reversible nature of H2RB binding and the development of tachyphylaxis limited their long-term efficacy[6]. In contrast, PPIs offer superior and sustained acid suppression, which has contributed to their widespread use and, in many cases, prolonged duration of therapy.

While the short-term safety and efficacy of PPIs are well established, concerns have emerged over potential long-term harms[7]. Observational studies have reported associations between prolonged PPI use and a variety of adverse outcomes, including chronic kidney disease[8], cardiovascular events[9], fractures[10], infections[11], and dementia[12]. Notably, several studies have also reported an increased risk of gastric cancer with long-term PPI use[13–20], particularly following *H. pylori* eradication[21]. Hypothesized mechanisms include PPI-induced hypergastrinemia[22] leading to mucosal proliferation[14], and alterations in the gastric microbiome[23] secondary to reduced acidity, potentially enabling carcinogenic processes[24].

However, the association between PPI use and gastric cancer remains controversial[25,26]. Critics have pointed out that patients prescribed PPIs often have underlying gastrointestinal symptoms or diseases—such as dyspepsia, reflux, or ulcers[1]—that themselves carry an increased risk of malignancy. In such settings, PPI use may not be causative, but rather a marker of early disease or its risk factors. Some studies have suggested that the observed association is explained by these underlying conditions, rather than by the drug itself[27]. Others have emphasized the importance of timing, noting that associations are strongest shortly before cancer diagnosis—raising the possibility of reverse causation or confounding by indication[28].

In this study, we evaluated the association between PPI use and upper gastrointestinal cancer using a matched case-control design. Importantly, we modelled drug exposure across multiple time windows prior to diagnosis, allowing us to test whether observed associations differ according to timing. We further adjusted for a range of gastrointestinal diagnoses that may reflect early cancer symptoms or predisposing conditions. By disentangling medication effects from underlying disease processes, we aimed to clarify whether PPIs independently contribute to cancer risk—or whether their association reflects confounding by indication or reverse causation.

## Methods

### Study Design and Data Source

We conducted a retrospective matched case-control study using electronic health records (EHRs) from Leumit Health Services (LHS), one of Israel’s four national health providers. LHS provides care to approximately 730,000 members and maintains a centralized longitudinal EHR system comprising over two decades of data on demographics, diagnoses, medication prescriptions and pharmacy purchases, laboratory tests, and healthcare utilization. All Israeli residents are entitled to universal healthcare under a standardized benefits package, and medication coverage is determined by a national formulary. Diagnoses in LHS are entered by treating physicians using International Classification of Diseases (ICD) codes. The accuracy of diagnostic coding in the LHS registry is considered high and has supported multiple publications in leading peer-reviewed journals[29,30].

### Study Population and Matching

The study included all LHS members with active enrolment between 2003 and 2024. Cases were defined as individuals aged 18–80 years with a diagnosis of upper gastrointestinal (GI) malignancy, identified through the Israeli National Cancer Registry and LHS diagnostic codes. The earliest cancer diagnosis date served as the index date. Individuals with a prior cancer diagnosis were excluded. Of the 892 detected upper GI cancer cases, 680 (76%) were identified via linkage to the Israeli National Cancer Registry, which provided structured data on tumour site, morphology, and histological grade (see Supplementary Tables 2-4). The remaining cases were identified through validated ICD-9 codes in the LHS medical record system (ICD-9 codes beginning with 150–152; Supplementary Table 5). Among registry-verified cases, the most common anatomical sites were gastric antrum (n = 93), cardia (n = 85), and body of stomach (n = 61). The most frequent morphologies were adenocarcinoma (n = 317), signet ring carcinoma (n = 104), and squamous cell carcinoma (n = 36). Poorly differentiated tumours (grade III) were the most prevalent (n = 275), followed by moderately differentiated tumours (grade II, n = 121).

Each case was matched to exactly ten cancer-free controls based on sex, ethnic sector (general population, Ultra-Orthodox Jewish, or Arab), socioeconomic status, and year of enrolment in LHS. Cases for which ten exact matches on these variables could not be identified were excluded. Among eligible controls, individuals with the closest possible date of birth and no history of cancer were selected.

### Sociodemographic Classification

Socioeconomic status was derived from residential address using the Points Location Intelligence® system, which ranks neighbourhoods on a 1–20 scale aligned with government indicators. Ethnic sector classification was based on residential patterns and demographic characteristics, using algorithms validated in previous population-based studies.

### Medication and Symptom Exposure Assessment

Medication exposures were determined from pharmacy purchase records covering up to 10 years prior to the index date. Medications were categorized using Anatomical Therapeutic Chemical (ATC) classification codes. In addition to proton pump inhibitors (PPIs), we included medications commonly used in our health system to treat upper gastrointestinal symptoms, namely famotidine (a histamine H2 receptor antagonist) and antacids (e.g., calcium carbonate). Exposure was assessed within predefined time windows: 0–0.1, 0.1–0.5, 0.5–1, 1–2, 2–3, and 3–10 years before the index date. For each drug and time window, exposure was defined as a binary variable, set to positive if at least one purchase occurred during the interval.

Upper gastrointestinal symptoms were identified from medical visits during the same 10-year period using ICD-9 diagnostic codes. Symptom categories included dyspepsia, gastroesophageal reflux, abdominal pain, gastritis, peptic ulcer disease, and *H. pylori* infection, with the list of ICD-9 codes detailed in Supplementary Table 6.

### Covariates

Additional covariates included age, smoking status, healthcare worker status, number of physician visits in the year before the index date, and pregnancy history (in women).

### Statistical Analyses

Differences between cases and controls were assessed using two-tailed t-tests for continuous variables and Fisher’s exact test for categorical variables. Conditional logistic regression was used to estimate adjusted odds ratios (aORs) for the association between medication use and upper GI malignancy, accounting for the matched design. Models were further adjusted for potential confounders. To address reverse causality and confounding by indication, we included time-binned medication exposures and constructed additional models excluding the year before diagnosis. Diagnostic codes for gastrointestinal conditions (e.g. gastritis, GERD, peptic ulcer disease) were included to control for symptomatic indications that may precede cancer diagnosis.

Analyses were performed using R version 4.4.0 (R Foundation for Statistical Computing). Data extraction and preprocessing were conducted using structured query language (T-SQL) and Python version 3.11 scripts developed by the Leumit Research Institute.

A two-sided P<0.05 was considered statistically significant.

### Patient and public involvement

As the study involved retrospective analysis of data, patients and members of the public were not directly involved in the study design.

## Results

Figure 1 illustrates the flowchart used to construct the study cohort. The study included 875 patients diagnosed with upper gastrointestinal (GI) cancer and 8,750 matched cancer-free controls (Table 1). Cases and controls were well matched by age, sex, ethnicity, and year of enrolment. The mean age in both groups was 63.0 years (standard deviation [SD] 11.9), and 37.5% were female. Ethnic distribution was matched by design, with 12.2% Arab, 10.3% Ultra-Orthodox Jewish, and 77.5% from the general population.

**Table 1:** Demographic and clinical characteristics of upper gastrointestinal cancer cases and matched controls.

|  |  | case | control | *P* value |
| --- | --- | --- | --- | --- |
| **N** |  | 875 | 8,750 |  |
| **Sex** | **Female** | 328 (37.5 %) | 3,280 (37.5 %) | 1.000 |
|  | **Male** | 547 (62.5 %) | 5,470 (62.5 %) | 1.000 |
| **Age (years)** |  | 63.0 ± 11.9 | 63.0 ± 11.9 | 0.999 |
| **BMI category** | **<18.5 Underweight** | 23 (3.03 %) | 87 (1.07 %) | <0.001 |
|  | **18.5-24.9 Normal** | 258 (34.04 %) | 2,181 (26.74 %) | <0.001 |
|  | **25-29.9 Overweight** | 281 (37.07 %) | 3,306 (40.53 %) | 0.063 |
|  | **≥30 Obese** | 196 (25.86 %) | 2,582 (31.66 %) | <0.001 |
| **BP systolic (mmHg)** |  | 129 ± 18 | 131 ± 21 | 0.003 |
| **BP diastolic (mmHg)** |  | 76.4 ± 9.9 | 77.7 ± 9.7 | <0.001 |
| **BP category** | **Hypertension 1** | 204 (23.9 %) | 2,135 (24.8 %) | 0.561 |
|  | **Hypertension 2** | 173 (20.3 %) | 2,043 (23.8 %) | 0.022 |
|  | **Normal BP** | 477 (55.9 %) | 4,417 (51.4 %) | 0.013 |
| **Smoking status** | **Non-smoker** | 508 (71.0 %) | 6,208 (76.8 %) | <0.001 |
|  | **Past smoker** | 27 (3.78 %) | 280 (3.46 %) | 0.670 |
|  | **Smoker** | 180 (25.2 %) | 1,595 (19.7 %) | <0.001 |
| **Ethnic group** | **Arab** | 107 (12.2 %) | 1,070 (12.2 %) | 1.000 |
|  | **General** | 678 (77.5 %) | 6,780 (77.5 %) | 1.000 |
|  | **Jewish Ultra-orthodox** | 90 (10.3 %) | 900 (10.3 %) | 1.000 |
| **Creatinine (mg/dL)** |  | 0.91 ± 0.44 | 0.91 ± 0.43 | 0.820 |
| **eGFR MDRD** | **(mL/min/1.73m<sup>2</sup>)** | 84.1 ± 24.5 | 83.4 ± 24.2 | 0.388 |
| **Triglycerides** | **(mg/dL)** | 129 ± 65 | 134 ± 77 | 0.039 |
| **Haemoglobin A1c** | **(%)** | 6.11 ± 1.07 | 6.16 ± 1.12 | 0.292 |
| **HDL Cholesterol** | **(mg/dL)** | 47.5 ± 13.0 | 49.7 ± 13.8 | <0.001 |
| **LDL Cholesterol** | **(mg/dL)** | 114 ± 36 | 115 ± 36 | 0.576 |
| **Macroalbuminuria** |  | 26 (2.97 %) | 229 (2.62 %) | 0.508 |
| **Microalbuminuria** |  | 112 (12.8 %) | 1,100 (12.6 %) | 0.831 |
| **Haemoglobin** | **(g/dL)** | 13.2 ± 2.0 | 14.0 ± 1.5 | <0.001 |

**Table 2:** Comparative analysis of medication exposure and gastrointestinal diagnoses recorded in the five years before the index date among upper gastrointestinal cancer cases and matched controls.

|  | Case (%) | Control (%) | Odds Ratio | P Value |
| --- | --- | --- | --- | --- |
| **Medication purchase in the last five years** |  |  |  |  |
| Famotidine PO | 139 (15.9%) | 1072 (12.3%) | 1.35 | 0.0027 |
| Omeprazole PO | 524 (59.9%) | 3175 (36.3%) | 2.62 | <0.0001 |
| Lansoprazole PO | 109 (12.5%) | 598 (6.8%) | 1.94 | <0.0001 |
| Esomeprazole PO | 210 (24.0%) | 739 (8.4%) | 3.42 | <0.0001 |
| Calcium Carbonate PO | 24 (2.7%) | 201 (2.3%) | 1.20 | 0.4104 |
| **Diagnosis recorded** |  |  |  |  |
| Peptic Ulcer Disease | 90 (10.3%) | 201 (2.3%) | 4.87 | <0.0001 |
| Abdominal pain | 607 (69.4%) | 3961 (45.3%) | 2.74 | <0.0001 |
| GERD | 186 (21.3%) | 1159 (13.2%) | 1.77 | <0.0001 |
| Gastritis | 181 (20.7%) | 864 (9.9%) | 2.38 | <0.0001 |
| *H. Pylori* | 187 (21.4%) | 954 (10.9%) | 2.22 | <0.0001 |

P values calculated using Fisher's exact test. Medication exposure reflects at least one pharmacy purchase in the five years preceding the index date. Diagnoses include any recorded occurrence in a ten-year window.

**Figure 1.**
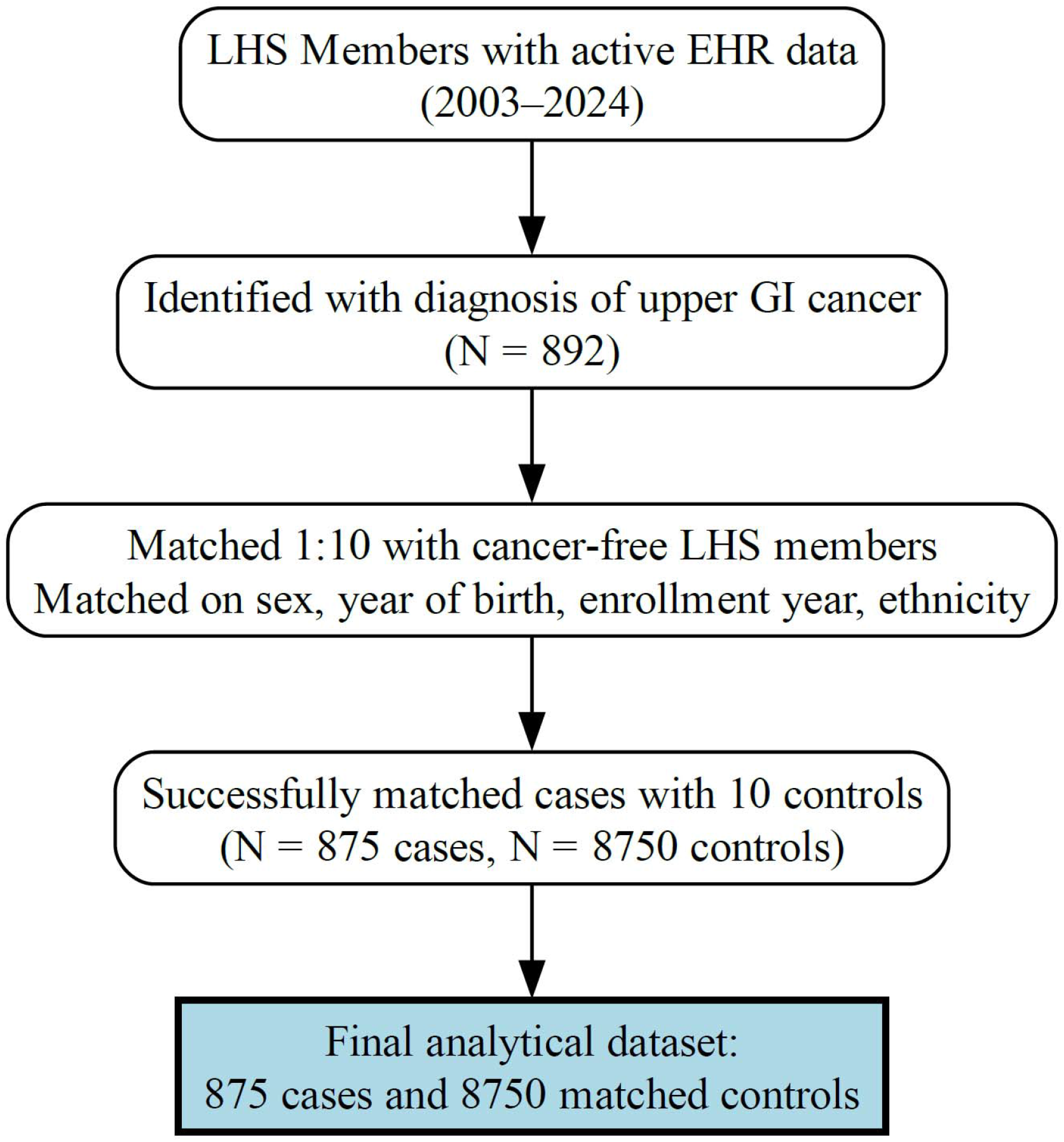
Flow diagram of the cohort

Cancer cases were more likely than controls to be underweight (3.0% vs. 1.1%) and less likely to be obese (25.9% vs. 31.7%). Current smoking was more common among cases (25.2% vs. 19.7%, p < 0.001), and the average socioeconomic status score was slightly lower (9.20 vs. 9.56, p = 0.004). Hypertension was slightly less prevalent in cases (20.3% vs. 23.8%, for stage 2 hypertension), and mean haemoglobin (13.2 vs. 14.0 g/dL) and HDL cholesterol (47.5 vs. 49.7 mg/dL) levels were lower among cases, though creatinine, LDL cholesterol, and HbA1c levels were similar between groups.

Medication exposure and gastrointestinal diagnoses in the years before the index date are summarized in Table 2. Proton pump inhibitor (PPI) use was significantly more common among cases than controls, including esomeprazole (24.0% vs. 8.4%; OR 3.42), omeprazole (59.9% vs. 36.3%; OR 2.62), and lansoprazole (12.5% vs. 6.8%; OR 1.94). Famotidine use was also more frequent among cases, while calcium carbonate showed no difference. Cancer cases were also more likely to have been diagnosed with upper GI conditions, including peptic ulcer disease, abdominal pain, gastritis, gastroesophageal reflux disease (GERD), and *H. pylori* infection. Multivariable conditional logistic regression models adjusting for demographic and clinical factors found that recent PPI use (within five years) was associated with increased odds of cancer (Figure 2A). Esomeprazole had the strongest association (adjusted OR 3.90, 95% CI 3.14 to 4.84), followed by omeprazole (aOR 2.60, 95% CI 2.22 to 3.06) and lansoprazole (aOR 1.42, 95% CI 1.12 to 1.81). Famotidine and calcium carbonate were not significantly associated with cancer risk.

**Figure 2.**
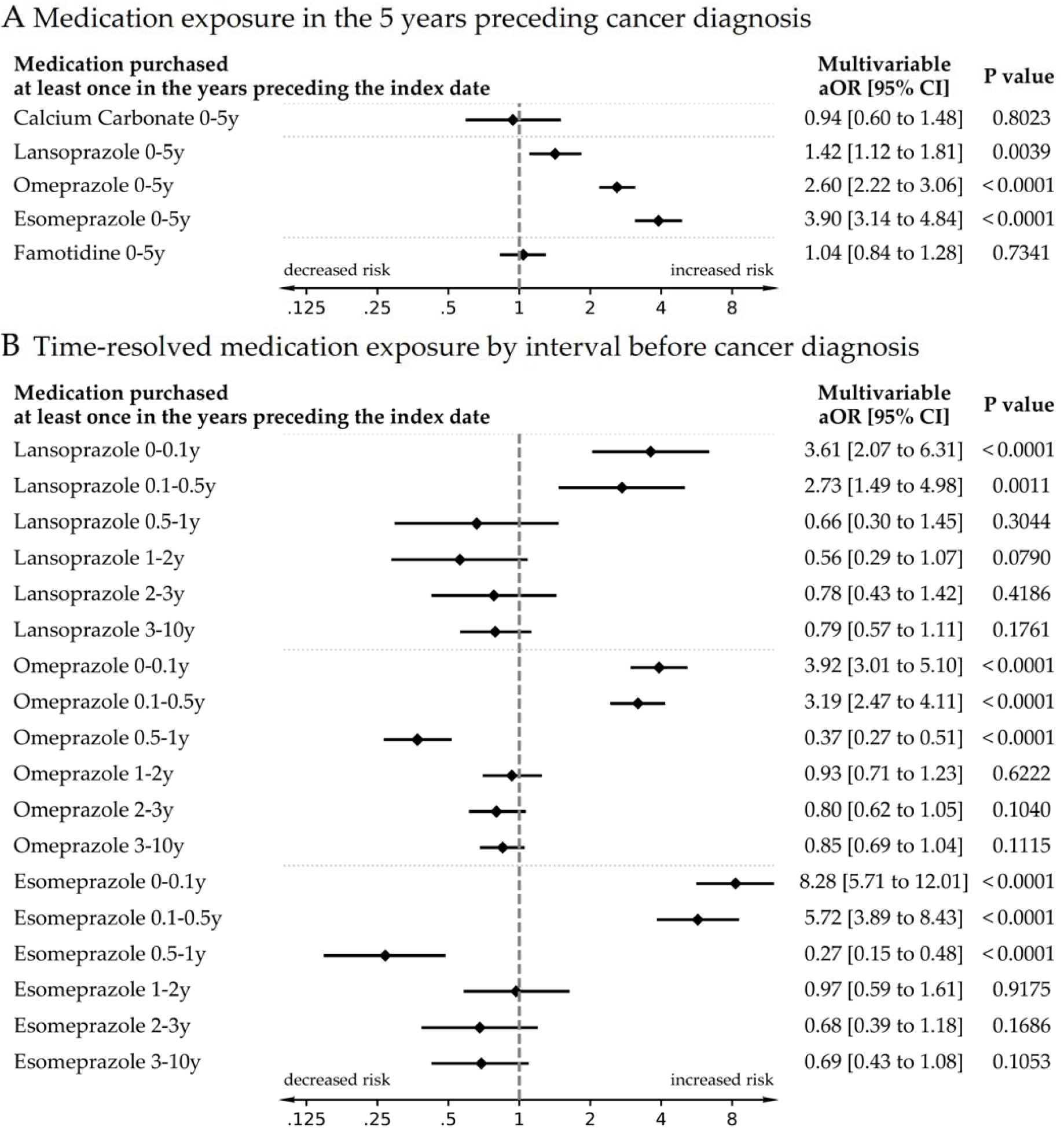
Association Between PPI Use and Upper Gastrointestinal Cancer: Impact of Exposure Timing. Forest plots illustrating the association between proton pump inhibitor (PPI) and related medication use and the risk of upper gastrointestinal (GI) cancer, based on multivariable conditional logistic regression models. **Panel A** shows odds ratios (ORs) for cancer associated with any medication exposure during the five years preceding diagnosis, adjusted for demographic and clinical covariates. **Panel B** presents time-resolved associations across multiple exposure windows, highlighting elevated risk during the final year before diagnosis and reduced or null risk for more distant exposures after adjustment for recent exposure.

In time-stratified analyses (Figure 2B), the highest risks were observed with PPI use within the year preceding cancer diagnosis, especially in the 0–0.1 year and 0.1–0.5 year windows. For esomeprazole, the adjusted odds ratios were 8.28 (95% CI 5.71 to 12.01) and 5.72 (95% CI 3.89 to 8.43), respectively. Risk declined with increasing time since last use, and no significant associations were found for PPI use more than one year before diagnosis.

In a model excluding PPI use during the final year and adjusting for GI diagnoses (Figure 3), no positive association remained between any PPI and cancer risk. Instead, exposure to omeprazole and lansoprazole in the 3–10 year window was associated with reduced odds of cancer (omeprazole: aOR 0.62, 95% CI 0.51 to 0.75; lansoprazole: aOR 0.68, 95% CI 0.49 to 0.94). Esomeprazole showed a similar but non-significant trend (aOR 0.68, 95% CI 0.45 to 1.01). Diagnoses such as peptic ulcer disease (aOR 3.70), abdominal pain (aOR 2.91), and GERD (aOR 1.66) remained strongly associated with cancer.

**Figure 3.**
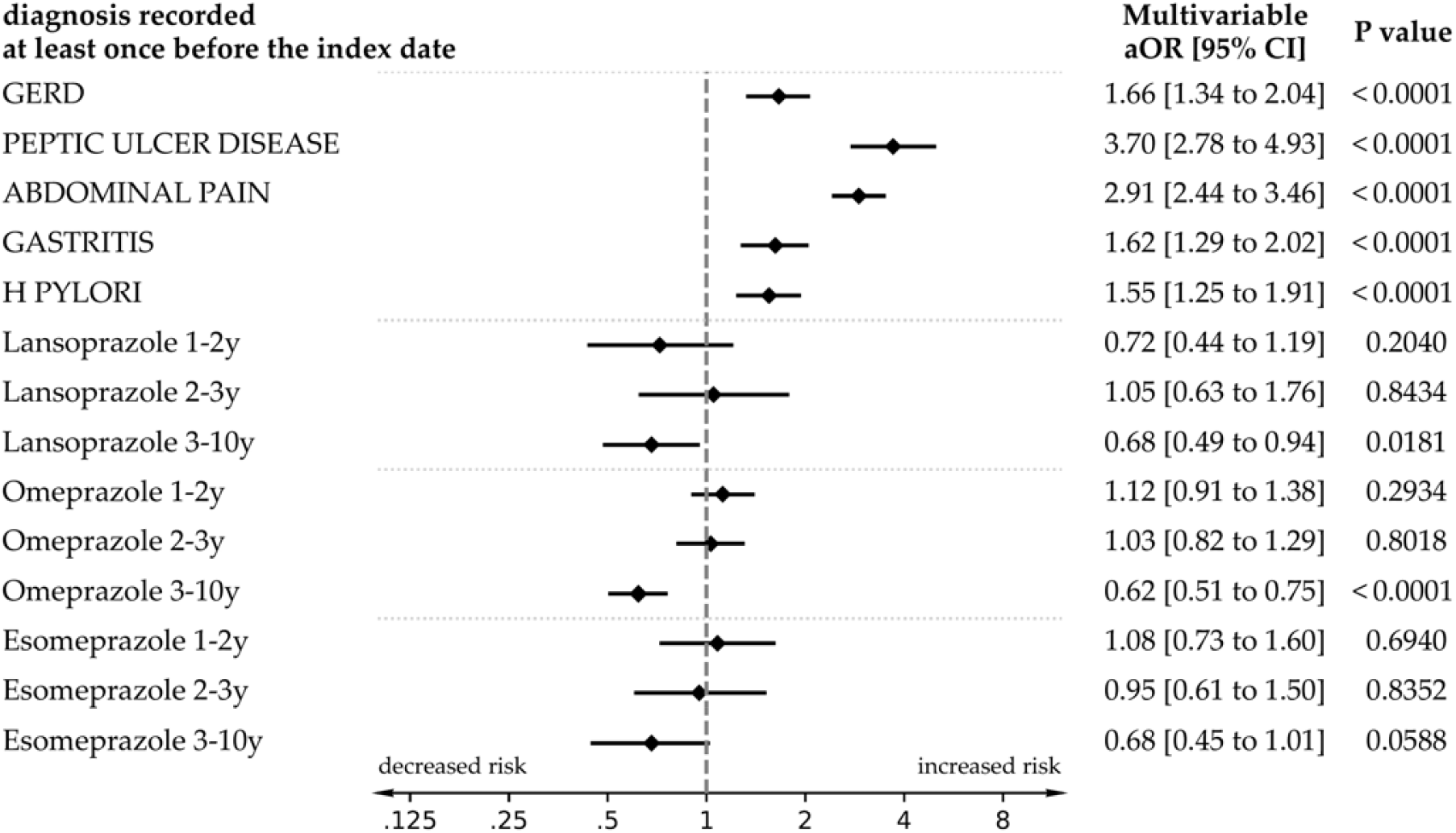
Association Between Past PPI Use and Upper Gastrointestinal Cancer Risk After Adjustment for Gastrointestinal Diagnoses. Forest plot showing adjusted odds ratios (aORs) and 95% confidence intervals for the association between proton pump inhibitor (PPI) use at different time windows prior to one year before the index date (1–2 years, 2–3 years, and 3–10 years) and the risk of upper gastrointestinal (GI) cancer. Models are adjusted for gastrointestinal diagnoses recorded at any time prior to the index date, including Helicobacter pylori infection, gastritis, peptic ulcer disease, abdominal pain, and gastroesophageal reflux disease (GERD). Time windows are treated as separate binary exposures, with medication exposure defined by ≥1 pharmacy purchase during the interval. The analysis uses multivariable conditional logistic regression within a matched case-control framework (1:10 matching). Findings highlight that after adjusting for underlying GI conditions, PPI use more than one year prior to cancer diagnosis is not associated with increased cancer risk—and in several cases is associated with decreased risk.

## Discussion

In this large matched case-control study based on electronic health records, we evaluated the association between proton pump inhibitor (PPI) use and the risk of upper gastrointestinal (GI) malignancies. While initial models indicated a strong association between PPI use—particularly esomeprazole and omeprazole—and upper gastrointestinal cancer risk, our temporally stratified and diagnosis-adjusted analyses showed that this association was largely driven by recent use. Once recent exposure was accounted for, associations with earlier use (beyond one year prior to diagnosis) lost statistical significance or appeared protective. In models that adjusted for GI symptoms, PPI use more than three years before cancer diagnosis was associated with a significantly reduced risk. These findings suggest that previously reported harmful associations may be explained by reverse causation and confounding by indication, rather than a true causal effect of long-term PPI therapy.

Reverse causation—also known as protopathic bias—is a form of bias that arises when a medication is prescribed for early symptoms of a disease that has not yet been diagnosed. In this context, early manifestations of a process associated with cancer risk (such as dyspepsia, reflux, or abdominal discomfort) may lead to the initiation of acid-suppressive therapy, creating a spurious association between the medication and subsequent cancer diagnosis. This can create a misleading association between the medication and the eventual cancer diagnosis. In our dataset, the adjusted odds of cancer were highest for PPI use in the 0–0.1 year and 0.1–0.5 year intervals before diagnosis, consistent with this mechanism. Many previous observational studies that reported increased cancer risk with PPI use did not account for timing of exposure[13–20] or for pre-existing gastrointestinal conditions that prompted treatment[1,28]. Consequently, they may have conflated symptomatic treatment with causative risk.

Our study addressed these limitations in several ways. First, we used a high-resolution, time-anchored dataset that allowed for temporal disaggregation of medication exposure windows. This made it possible to identify that only recent PPI use was associated with increased risk, while more remote exposure had no such association—or even appeared protective after accounting for recent exposure. Second, we adjusted for a comprehensive set of GI diagnoses frequently preceding PPI use, including gastritis, GERD, peptic ulcer disease, abdominal pain, and *H. pylori* infection. When these clinical indications were included in the model and recent medication exposure was excluded, the association between PPI use and cancer disappeared or reversed, with PPIs (especially omeprazole and lansoprazole) showing a statistically significant protective association.

These findings also align with biological plausibility. Chronic mucosal injury—such as that caused by persistent acid reflux or untreated *H. pylori* infection—is a well-established risk factor for gastric and oesophageal adenocarcinoma[31]. PPIs reduce gastric acid secretion, promote mucosal healing, and are effective in mitigating these precancerous processes[32]. Prior studies have shown that PPI therapy in patients with Barrett’s oesophagus or chronic gastritis may reduce progression to dysplasia or malignancy[33,34]. The observed protective association with long-term PPI use in our study could reflect effective acid suppression and mucosal stabilization in patients at elevated baseline risk.

An additional consideration is the interaction between PPI therapy and *H. pylori* infection. PPIs are an integral part of *H. pylori* eradication regimens, which also include antibiotic therapy. Successful eradication[35], followed by mucosal healing under acid suppression, reduces the risk of atrophic gastritis and intestinal metaplasia. However, when *H. pylori* persists—particularly under prolonged acid suppression—gastrin levels rise, and altered gastric pH may allow the pathogen to persist in an inflammatory niche[23]. Thus, the net effect of long-term PPI use may vary depending on whether *H. pylori* has been eradicated or remains untreated. Our data could not directly assess *H. pylori* treatment success, but the protective associations seen with remote PPI use may reflect earlier eradication or stabilization of chronic inflammation.

This study has several strengths, including a large sample size; comprehensive electronic health records for over two decades documenting pharmacy purchases and ICD-coded diagnoses at each clinical encounter; precise malignancy onset dating that enabled rigorously timed case-control matching; robust adjustment for potential confounders; and high-resolution temporal analysis of medication exposure. Nonetheless, certain limitations warrant consideration. Residual confounding may persist, particularly due to unmeasured variables such as dietary patterns, or adherence to *H. pylori* eradication protocols. While ICD-coded diagnoses were assigned by treating physicians at the time of encounter— minimizing misclassification and recall bias—some clinically relevant factors, such as symptom severity or undocumented conditions, may not be fully captured within structured EHR data fields.

## Conclusions

In conclusion, our findings do not support a causal link between long-term PPI use and upper GI cancer. Rather, the observed associations in previous studies likely reflect reverse causation and confounding by clinical symptoms that precede diagnosis. When appropriately adjusted for diagnostic context and timing, PPI use is not associated with increased cancer risk and may be associated with lower risk when used in the more distant past.

## Data Availability

Because this study is based on individual-level clinical data, data sharing is restricted. Access to the data may be granted to qualified researchers upon reasonable request, subject to approval by the Leumit Health Services Institutional Review Board (IRB).

## Footnotes

### Statement

The Corresponding Author has the right to grant on behalf of all authors and does grant on behalf of all authors, an exclusive licence (or non exclusive for government employees) on a worldwide basis to the BMJ Publishing Group Ltd to permit this article (if accepted) to be published in BMJ editions and any other BMJPGL products and sublicences such use and exploit all subsidiary rights, as set out in our licence.

### Contributors

All authors provided final approval to publish. AI, EM had access to the raw data. AI and AOS designed the study. AI, IOS, ZD, EG, IG, EM, SV, AOS contributed to data analysis and interpretation. AI, IOS and AOS contributed to the drafting of the article. The corresponding author attests that all listed authors meet authorship criteria and that no others meeting the criteria have been omitted. AI is the guarantor.

### Competing interests

All authors have completed the ICMJE uniform disclosure form at www.icmje.org/coi_disclosure.pdf. The authors declare no competing interests. AI, ZD, IG, EM, and SV are employees of Leumit Health Services. All authors declare that they have no other relationships or activities that could appears to have influenced the submitted work.

### Funding

This research was internally funded by Leumit Health Services and was supported in part by the Intramural Research Program, National Institutes of Health, National Cancer Institute, Center for Cancer Research. The funders had no role in considering the study design or in the collection, analysis, interpretation of data, writing of the report, or decision to submit the article for publication. The content of this publication does not necessarily reflect the views or policies of the Department of Health and Human Services, nor does mention of trade names, commercial products, or organisations imply endorsement by the US government.

### Ethical approval

This study was approved by the Leumit Health Services Institutional Review Board (IRB) with a waiver of informed consent (approval number: LEU-0010-21). The waiver was justified on the basis that this large retrospective study used de-identified clinical data and posed no risk to participants.

### Transparency

The manuscript’s guarantor (AI) affirms that this manuscript is an honest, accurate, and transparent account of the study being reported; that no important aspects of the study have been omitted; and that any discrepancies from the study as planned have been explained.

### Source code sharing

The statistical analysis was performed using R. While the code has not been publicly deposited, it is available upon reasonable request from the corresponding author.

http://creativecommons.org/licenses/by-nc/4.0/

This is an Open Access article distributed in accordance with the Creative Commons Attribution Non Commercial (CC BY-NC 4.0) license, which permits others to distribute, remix, adapt, build upon this work non-commercially, and license their derivative works on different terms, provided the original work is properly cited and the use is non-commercial. See: http://creativecommons.org/licenses/by-nc4.0/.

## Notes

### Competing Interest Statement

The authors have declared no competing interest.

## References

1 Freedberg DE, Kim LS, Yang Y-X. The Risks and Benefits of Long-term Use of Proton Pump Inhibitors: Expert Review and Best Practice Advice From the American Gastroenterological Association. Gastroenterology. 2017;152:706–15. doi: 10.1053/j.gastro.2017.01.031

2 Strand DS, Kim D, Peura DA. 25 Years of Proton Pump Inhibitors: A Comprehensive Review. Gut Liver. 2017;11:27–37. doi: 10.5009/gnl15502

3 Schunack W. Pharmacology of H2-receptor antagonists: an overview. J Int Med Res. 1989;17 Suppl 1:9A–16A.

4 Shin JM, Sachs G. Pharmacology of proton pump inhibitors. Curr Gastroenterol Rep. 2008;10:528–34. doi: 10.1007/s11894-008-0098-4

5 MacLaren R, Reynolds PM, Allen RR. Histamine-2 Receptor Antagonists vs Proton Pump Inhibitors on Gastrointestinal Tract Hemorrhage and Infectious Complications in the Intensive Care Unit. JAMA Intern Med. 2014;174:564. doi: 10.1001/jamainternmed.2013.14673

6 Lilly CM, Aljawadi M, Badawi O, et al. Comparative Effectiveness of Proton Pump Inhibitors vs Histamine Type 2 Receptor Blockers for Preventing Clinically Important Gastrointestinal Bleeding During Intensive Care. Chest. 2018;154:557–66. doi: 10.1016/j.chest.2018.05.015

7 Maes ML, Fixen DR, Linnebur SA. Adverse effects of proton-pump inhibitor use in older adults: a review of the evidence. Ther Adv Drug Saf. 2017;8:273–97. doi: 10.1177/2042098617715381

8 Xie Y, Bowe B, Li T, et al. Proton Pump Inhibitors and Risk of Incident CKD and Progression to ESRD. Journal of the American Society of Nephrology. 2016;27:3153–63. doi: 10.1681/ASN.2015121377

9 Duarte GJ, Lopez J, Sosa F, et al. Proton pump inhibitors and cardiovascular risk: a critical review. Future Cardiol. 2024;20:779–94. doi: 10.1080/14796678.2024.2412910

10 Targownik LE, Lix LM, Metge CJ, et al. Use of proton pump inhibitors and risk of osteoporosis-related fractures. Can Med Assoc J. 2008;179:319–26. doi: 10.1503/cmaj.071330

11 Gulmez SE. Use of Proton Pump Inhibitors and the Risk of Community-Acquired Pneumonia. Arch Intern Med. 2007;167:950. doi: 10.1001/archinte.167.9.950

12 Gomm W, von Holt K, Thomé F, et al. Association of Proton Pump Inhibitors With Risk of Dementia. JAMA Neurol. 2016;73:410. doi: 10.1001/jamaneurol.2015.4791

13 Abrahami D, McDonald EG, Schnitzer ME, et al. Proton pump inhibitors and risk of gastric cancer: population-based cohort study. Gut. 2022;71:16–24. doi: 10.1136/gutjnl-2021-325097

14 Cheung KS, Leung WK. Long-term use of proton-pump inhibitors and risk of gastric cancer: a review of the current evidence. Therap Adv Gastroenterol. 2019;12. doi: 10.1177/1756284819834511

15 Joo MK, Park J-J, Chun HJ. Proton pump inhibitor: The dual role in gastric cancer. World J Gastroenterol. 2019;25:2058–70. doi: 10.3748/wjg.v25.i17.2058

16 Peng T-R, Wu T-W, Li C-H. Association between proton-pump inhibitors and the risk of gastric cancer: a systematic review and meta-analysis. Int J Clin Oncol. 2023;28:99–109. doi: 10.1007/s10147-022-02253-2

17 Waldum HL, Sørdal Ø, Fossmark R. Proton pump inhibitors (PPIs) may cause gastric cancer – clinical consequences. Scand J Gastroenterol. 2018;53:639–42. doi: 10.1080/00365521.2018.1450442

18 Segna D, Brusselaers N, Glaus D, et al. Association between proton-pump inhibitors and the risk of gastric cancer: a systematic review with meta-analysis. Therap Adv Gastroenterol. 2021;14. doi: 10.1177/17562848211051463

19 Ko Y, Tang J, Sanagapalli S, et al. Safety of proton pump inhibitors and risk of gastric cancers: review of literature and pathophysiological mechanisms. Expert Opin Drug Saf. 2016;15:53–63. doi: 10.1517/14740338.2016.1118050

20 Xia B, He Q, Smith FG, et al. Individualized prevention of proton pump inhibitor related adverse events by risk stratification. Nat Commun. 2024;15:3591. doi: 10.1038/s41467-024-48007-8

21 Cheung KS, Chan EW, Wong AYS, et al. Long-term proton pump inhibitors and risk of gastric cancer development after treatment for Helicobacter pyloriJ: a population-based study. Gut. 2018;67:28–35. doi: 10.1136/gutjnl-2017-314605

22 Przemeck SMC, Varro A, Berry D, et al. Hypergastrinemia increases gastric epithelial susceptibility to apoptosis. Regul Pept. 2008;146:147–56. doi: 10.1016/j.regpep.2007.09.002

23 Imhann F, Bonder MJ, Vich Vila A, et al. Proton pump inhibitors affect the gut microbiome. Gut. 2016;65:740–8. doi: 10.1136/gutjnl-2015-310376

24 Parsons BN, Ijaz UZ, D’Amore R, et al. Comparison of the human gastric microbiota in hypochlorhydric states arising as a result of Helicobacter pylori-induced atrophic gastritis, autoimmune atrophic gastritis and proton pump inhibitor use. PLoS Pathog. 2017;13:e1006653. doi: 10.1371/journal.ppat.1006653

25 Vaezi MF, Yang Y-X, Howden CW. Complications of Proton Pump Inhibitor Therapy. Gastroenterology. 2017;153:35–48. doi: 10.1053/j.gastro.2017.04.047

26 Robertson DJ, Larsson H, Friis S, et al. Proton Pump Inhibitor Use and Risk of Colorectal Cancer: A Population-Based, Case–Control Study. Gastroenterology. 2007;133:755–60. doi: 10.1053/j.gastro.2007.06.014

27 Moayyedi P, Veldhuyzen van Zanten SJO, Hookey L, et al. Proton pump inhibitors and gastric cancer: association is not causation. Gut. 2019;68:1529.2-1530. doi: 10.1136/gutjnl-2018-316958

28 Crafa P, Franceschi M, Rodriguez-Castro KI, et al. PPIs and gastric cancer: any causal relationship? Acta Biomed. 2023;94:e2023096. doi: 10.23750/abm.v94i3.14105

29 Israel A, Merzon E, Schäffer AA, et al. Elapsed time since BNT162b2 vaccine and risk of SARS-CoV-2 infection: test negative design study. The BMJ. 2021;375:e067873. doi: 10.1136/bmj-2021-067873

30 Israel A, Raz I, Vinker S, et al. Type 2 Diabetes in Patients with G6PD Deficiency. New England Journal of Medicine. 2024;391:568–9. doi: 10.1056/NEJMc2406156

31 Uemura N, Okamoto S, Yamamoto S, et al. Helicobacter pylori Infection and the Development of Gastric Cancer. New England Journal of Medicine. 2001;345:784–9. doi: 10.1056/NEJMoa001999

32 Wolfe MM, Sachs G. Acid suppression: Optimizing therapy for gastroduodenal ulcer healing, gastroesophageal reflux disease, and stress-related erosive syndrome. Gastroenterology. 2000;118:S9–31. doi: 10.1016/S0016-5085(00)70004-7

33 Conteduca V, Sansonno D, Ingravallo G, et al. Barrett’s esophagus and esophageal cancer: An overview. Int J Oncol. 2012;41:414–24. doi: 10.3892/ijo.2012.1481

34 Chen Y, Sun C, Wu Y, et al. Do proton pump inhibitors prevent Barrett’s esophagus progression to high-grade dysplasia and esophageal adenocarcinoma? An updated meta-analysis. J Cancer Res Clin Oncol. 2021;147:2681–91. doi: 10.1007/s00432-021-03544-3

35 Suzuki H, Nishizawa T, Hibi T. Helicobacter Pylori Eradication Therapy. Future Microbiol. 2010;5:639–48. doi: 10.2217/fmb.10.25

